# Prevalence and Practice of Traditional Uvulectomy Among Sudanese Children and Adolescents, Along with Maternal Perception and Associated Risk Factors: A Community-Based Cross-Sectional Study

**DOI:** 10.1101/2025.07.03.25330812

**Authors:** Jaber Hamad Jaber Amin, Ahmed Alshafei Elmahi Ahmed, Mohamedelmustafa Yahya Mohamed Eldouma, Alshafee Mohammed Jebreel Sulyman, Amna A. Eltayeb, Asjad Zainalabdeen Ahmed, Huyam Elsir Fadlelseed Babiker, Tanzeel Mohamedain Abuelgasim Abdalla, Dania A. Elsiddig, Majdy Jailany Alamin Abdelgadir, Nooralsham Abdalla Adam Yousif, Israa Osman Ali Alhassan, Zainab Salih Ahmed Adam, Mohammed Hammad Jaber Amin

## Abstract

**Background:** Traditional uvulectomy (TU) is the removal of the uvula, either partially or totally by the traditional healer. It is prevalent in sub-Saharan Africa, driven by cultural beliefs and perceived therapeutic benefits, despite the risks of complications -bleeding, infections, and death, and limited data exist. Our study aimed to assess the prevalence and practice of traditional uvulectomy among Sudanese children and adolescents, along with maternal perceptions, and identify associated risk factors.

**Method:** A descriptive cross-sectional community-based study (from January to April 2025) involved 1,135 mothers of children ≤18 years, across more than ten states. Data were collected via face-to-face interviews using a validated structured questionnaire to assess the socio-demographics, TU perception, practice, and complications. A convenience sampling technique was used. We analyzed with R software, and p < 0.05 was considered statistically significant.

**Results:** The prevalence was 15%, peaking in infants (<6 years: 16%) and adolescents (12–18 years: 17%). Maternal belief was significantly associated with younger child age (37% for 12–18 years vs 16% for 6–12 years; p<0.001). The reasons included breastfeeding difficulties (18%), cultural tradition (16%), and failure to thrive (13%). Mothers (45%) and grandmothers (41%) primarily decided to perform TU, with traditional healers conducting 55% of procedures using unsterilized tools. Complications included fever (35%), feeding difficulties (21%), and bleeding (13%). TU was associated with younger maternal age (median 22 years), lower education (21% illiterate), and tribal tradition (92% adherence), all p<0.001.

**Conclusion:** TU persists in Sudan due to cultural entrenchment, intergenerational influence, and healthcare barriers. Interventions must engage tribal leaders and healers, promote safer alternatives, and improve education and healthcare.

## Introduction

Traditional uvulectomy (TU) is the removal of the uvula partially or totally by a traditional healer. The uvula is a small, fleshy lobe that is present above the throat, between the two lymphoid tissues or tonsils, and descends from the palate at the lower central border. [1,2] It plays a key role in various physiological functions, particularly in moisturizing oropharyngeal mucosa, serving for linguistic communication, and increasing immunologic function by discharging small amounts of saliva of high antibody activity, also by conjunction with the soft palate closing the nasopharynx, therefore, preventing aspiration and regurgitation of food or liquid into the nasal cavity [3] It develops from soft palate by fusion of the two parts at the 11th week. It has a muscle called the musculus uvulae, which shortens after contraction. The arterial supply comes primarily from the ascending pharyngeal, the ascending palatine, and the descending palatine arteries, while venous drainage flows through the pterygoid plexus into the internal jugular vein. The lymphatic drainage is principally to the upper deep cervical group of nodes, the mucous membrane is innervated by the lesser palatine and glossopharyngeal nerves, while the muscles are supplied by the pharyngeal plexus [4-6].

Traditional healers in many communities are trusted providers, using natural remedies and cultural procedures to treat ailments [7]. Traditional African medicine, like traditional uvulectomy, passed down through generations, included practices [8]. TU is rarely performed in hospitals, except in procedures like uvulopalatoplasty and adenotonsillectomy [9,10]. TU is widely practiced in Sub-Saharan Africa, Nigeria, Kenya, Sierra Leone, Tanzania, Ethiopia, South Africa, and Sudan [11,12-16,17]. it is also prevalent in Israel, Saudi Arabia, and several Middle Eastern countries [11,18,19]. Cultural beliefs, family traditions, and socio-economic factors perpetuate the practice, for example, 33 % of mothers practiced TU, and 25.7 % were due to family tradition [17]. Low socio-demographic status, lack of education, and limited access to healthcare further drive its persistence [20,21]

The rationale for uvulectomy varies; parents fear sudden death from an obstructed airway, and throat issues in childhood, like soft swallowing and hoarseness, recurrent throat infections, failure to thrive, recurrent fever, feed aversion, and chronic cough [3,20]. male children, often seen as a source of family pride, are more likely to receive immediate traditional care, including uvulectomy, when ill [3]. The procedure is done using unsterilized knives, without anesthesia, and herbs are applied to the wound [22]. Severe complications included severe bleeding, anemia, hepatitis, infection, tetanus, HIV risk, abscesses, aspiration, and consequent upper airway obstruction and death [9,23,24]. other risks included jaundice, throat burn, prolonged pain, voice changes, sleep disturbances, breast milk regurgitation, and cavernous sinus thrombosis. [25] These complications lead to higher healthcare use, including antibiotics, oxygen therapy, IV fluids, blood transfusions, and phototherapy, escalating healthcare costs [3,21,23]. Uvulectomy-related complications contribute significantly to under-five childhood mortality. In Ethiopia, 20% of children undergo TU, with 15% experiencing severe complications requiring hospitalization [22] A qualitative study in Ethiopia identified cultural beliefs, access to traditional healers, family pressure, lack of awareness of uvulectomy benefits, and distance from healthcare facilities as primary drivers of uvulectomy [21]. In Sudan, 17.9 % of children under five years undergo it, with maternal age, education level, and regional background independently associated with the practice [13]. This study aims to assess the prevalence and practice of traditional uvulectomy among Sudanese children and adolescents, along with maternal perceptions about this practice, and identify associated risk factors. The findings will inform policy decisions and health programs to address TU to ensure child safety and correct health-seeking behavior.

## Methodology

This is a descriptive cross-sectional community-based study that assesses the prevalence and practice of traditional uvulectomy (TU) among Sudanese children and adolescents, along with perceptions of mothers regarding this practice, from 1^st^ January to 20^th^ April, 2025. We interviewed 1,135 mothers in more than ten states, and included all mothers with at least one child or adolescent less than 18 years old who agreed to participate in our study; declining participation or those with children outside the age range were excluded. A convenience sampling technique was used. The data were collected by trained medical students, who interviewed the mothers, face-to-face, in a private setting using a structured questionnaire adapted from validated previous studies. It was structured in English and translated into local languages to ensure clarity, then back-translated to ensure accuracy. The interviewers clarified each question before recording the answer. They trained to assess the socio-demographics (age, education, tribe), knowledge/perceptions regarding TU, practice information (tool, practitioner, timing), and complications. To decrease the bias, interviewers clarified each question without leading participants, and the data collection process is strictly supervised by a senior researcher.

Statistical analysis was performed using R software (version 4.4.2) utilizing tidyverse and tidyverse-associated packages, and p < 0.05 was considered statistically significant. Categorical variables were summarized using frequency percentages and compared using Chi-squared test or Fisher’s exact test. Continuous variables were summarized using medians and interquartile ranges and compared using the Wilcoxon rank sum test.

## Ethical approval

Ethical approval for the study was obtained from the general administration of health systems and research at the federal ministry of health, River Nile state, Sudan. The study was conducted in accordance with the principles of the Declaration of Helsinki. Oral consent was obtained from all participants. We were assured of confidentiality through anonymization of the data, and mothers retained the right to withdraw without consequences.

## Results

The study includes 1,135 mothers with a median age of 28, 38% of whom held graduate degrees. They typically had three children, with the youngest child often aged 1–3 years (35%). Additionally, most (61%) reported moderate income levels (Table 1).

**Table 1:**
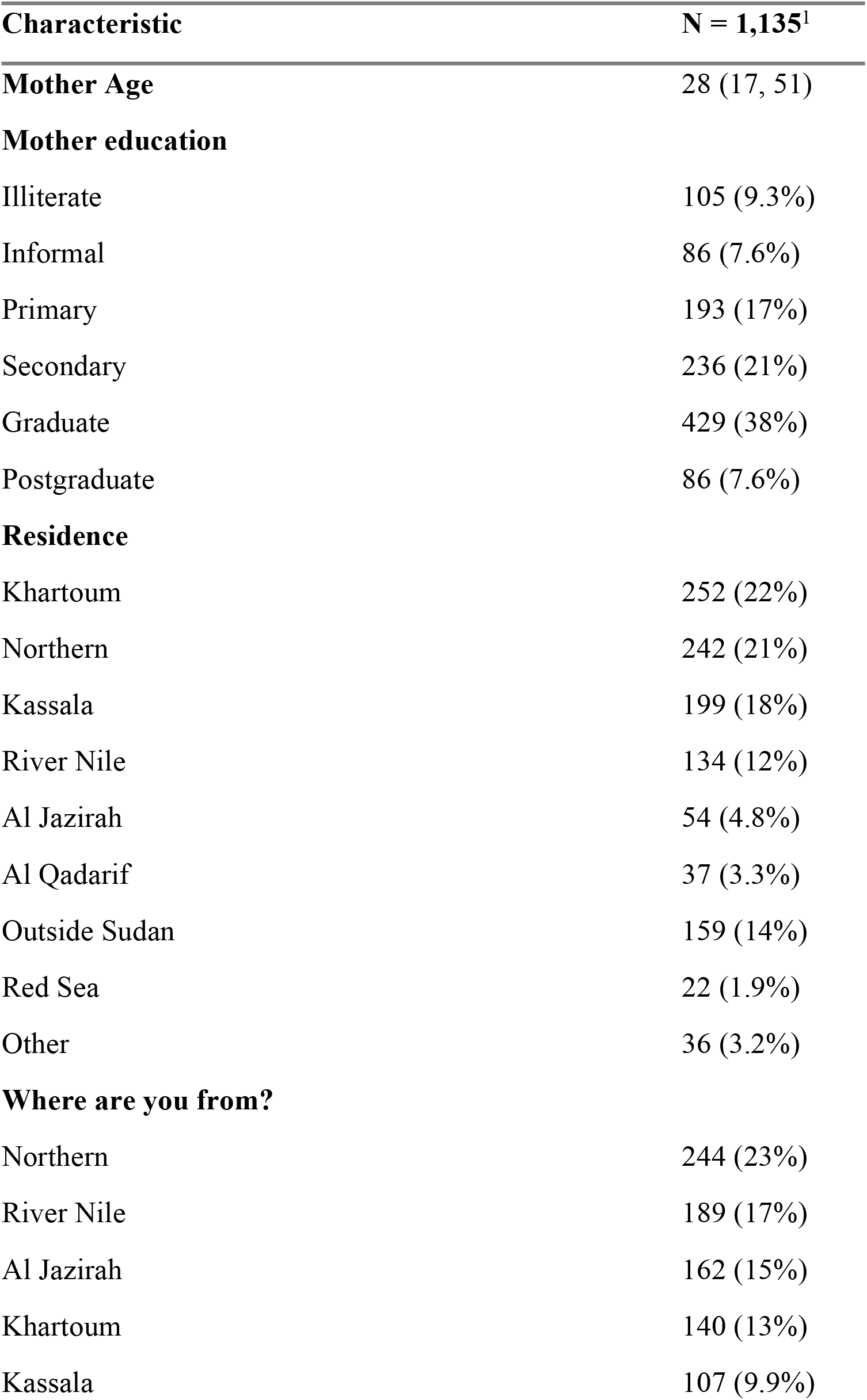
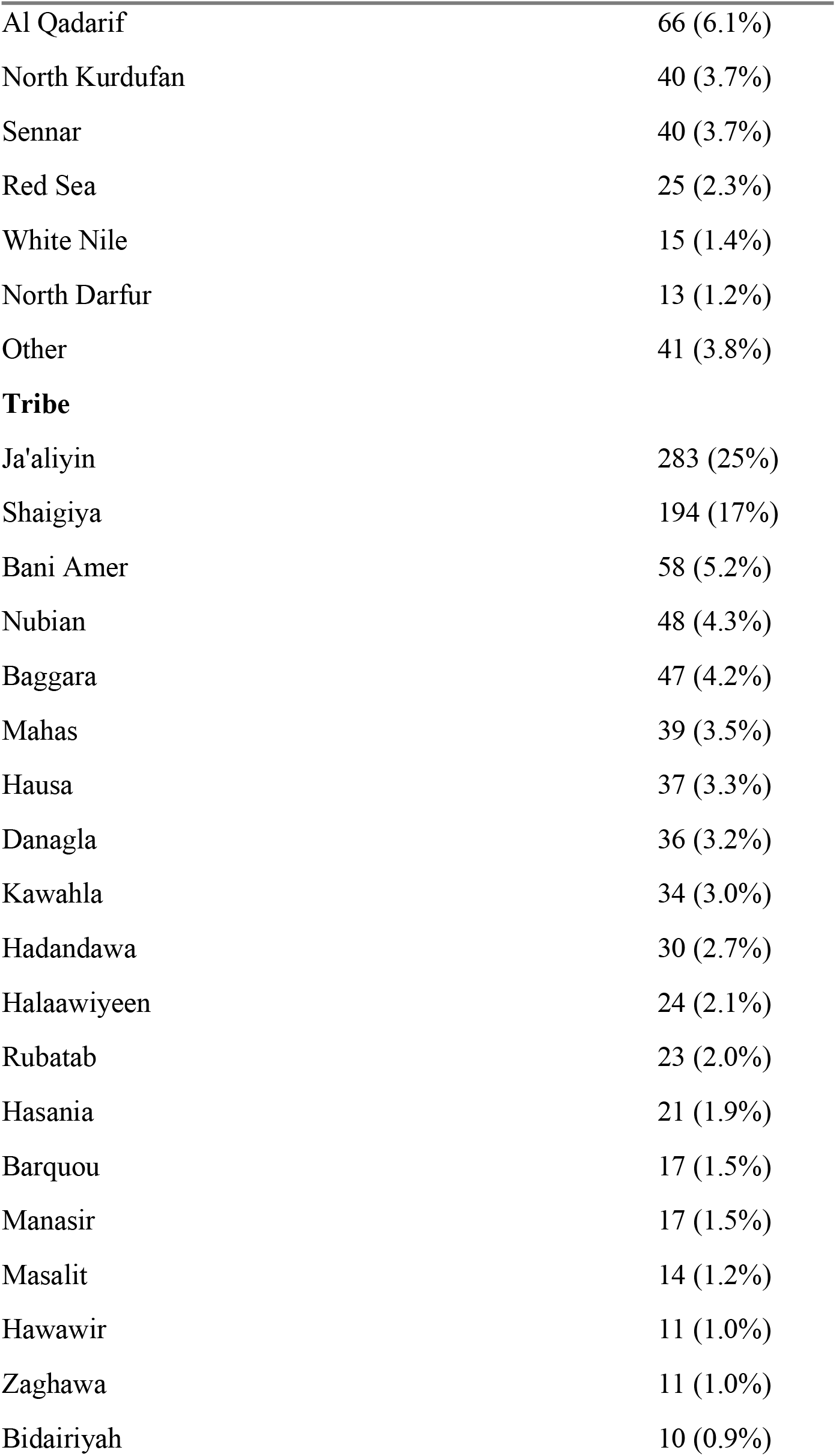
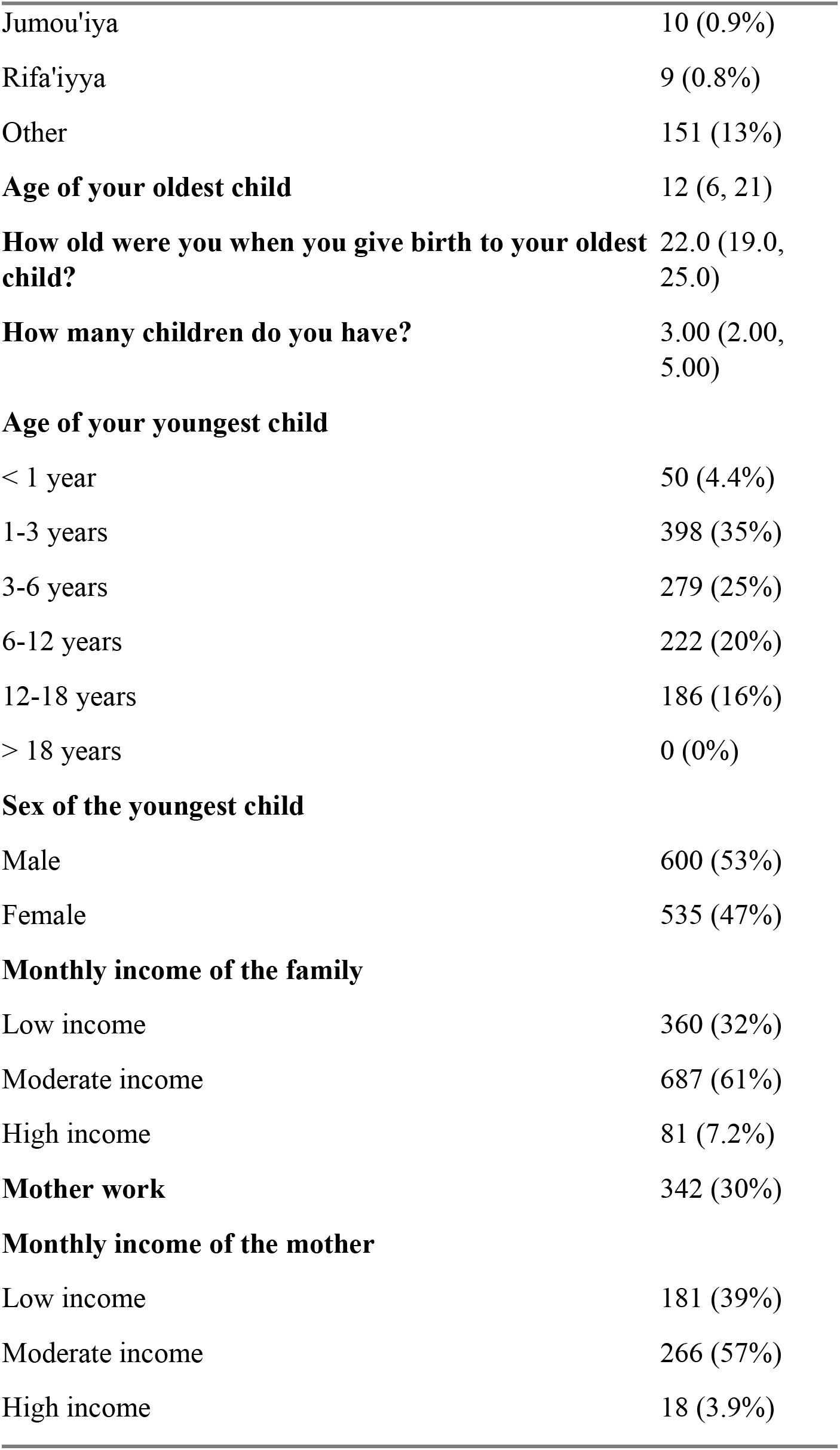
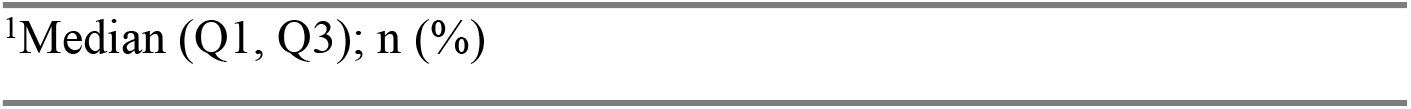
Sociodemographic characteristics of the participants.

### Prevalence and Perception

About 57% of mothers were aware of uvulectomy, with 35% acknowledging its presence in their tribe. Belief in the practice differed significantly by the youngest child’s age: 37% of mothers with children aged 12–18 years endorsed it, compared to 16% in the 6–12 years group (p<0.001). Similarly, 17% of mothers with children under six intended to schedule the procedure, compared to 6.2% for older children (p<0.001). Overall, 15% of children underwent uvulectomy, with higher rates in younger age groups (16% for <6 years vs. 17% for 12–18 years; p=0.020) (Table 2).

**Table 2:** Prevalence and perception of uvulectomy practice among the mothers divided by their youngest child age.

| Characteristic | Overall<br>N =<br>1,135<sup>1</sup> | < 6<br>years<br>N =<br>727<sup>1</sup> | 6-12<br>years<br>N = 222<sup>1</sup> | 12-18<br>years<br>N = 186<sup>1</sup> | p-<br>value<sup>2</sup> |
| --- | --- | --- | --- | --- | --- |
| **Have you heard of the traditional uvulectomy practice in children?** | 652<br>(57%) | 407<br>(56%) | 126<br>(57%) | 119<br>(64%) | 0.14 |
| **Presence of uvulectomy practice in your tribe** | 365<br>(35%) | 230<br>(33%) | 74 (38%) | 61 (40%) | 0.2 |
| **Do you totally believe in these practices?** | 206<br>(24%) | 139<br>(24%) | 25 (16%) | 42 (37%) | <0.001 |
| **Will you schedule your child for uvulectomy?** | 158<br>(14%) | 120<br>(17%) | 13<br>(6.2%) | 25 (14%) | <0.001 |
| **Did your child have his/her uvula removed** | 165<br>(15%) | 115<br>(16%) | 19<br>(8.7%) | 31 (17%) | 0.020 |

<sup>1</sup>n (%)
<sup>2</sup>Pearson's Chi-squared test

### Reasons for Uvulectomy

The most common reasons were problems with breastfeeding (18%), tradition (16%), and failure to gain weight (13%). Surprisingly, 30% of the mothers found uvulectomy effective, whereas 36% were undecided. Notably, 24% feared harm if the uvula was not cut, whereas 47% disagreed (Table 3).

**Table 3:** Reasons for performing uvulectomy among the participants.

| **Characteristic** | **N =**<br>**1,135<sup>1</sup>** |
| --- | --- |
| **Reasons for uvulectomy practice** |  |
| - **Difficulty breastfeeding** | 205<br>(18%) |
| - **Cultural reasons/ Family tradition** | 176<br>(16%) |
| - **Failure to gain weight** | 147<br>(13%) |
| - **Vomiting** | 144<br>(13%) |
| - **Loss of appetite** | 137<br>(12%) |
| - **Cough** | 82 (7.2%) |
| - **Preventive** | 60 (5.3%) |
| - **Fever** | 22 (1.9%) |
| - **Diarrhea** | 24 (2.1%) |
| - **Other** | 32 (2.8%) |
| **If other reasons, please specify** |  |
| Difficulty breathing-snoring | 11 (31%) |
| Sore throat | 4 (11%) |
| Inflammation of uvula | 3 (8.3%) |
| Chronic cough | 3 (8.3%) |
| Difficulty breathing-hearing problems | 3 (8.3%) |
| Tonsillitis | 2 (5.6%) |
| Recurrent fever | 1 (2.8%) |
| Delay in speech | 1 (2.8%) |
| Difficulty swallowing | 1 (2.8%) |
| Halitosis | 1 (2.8%) |
| Sunken eyes | 1 (2.8%) |
| Headache | 1 (2.8%) |
| Thirst | 1 (2.8%) |
| **Is uvulectomy a good treatment for the above?** |  |
| No | 278<br>(34%) |
| Not sure | 302<br>(36%) |
| Yes | 248<br>(30%) |
| **Does the uvula cause oropharyngeal blockage in children?** |  |
| No | 296<br>(37%) |
| Not sure | 297<br>(37%) |
| Yes | 209<br>(26%) |
| **If the uvula is not removed, will their presence harm the child?** |  |
| No | 395<br>(47%) |
| Not sure | 237<br>(28%) |
| Yes | 202<br>(24%) |

| Characteristic | N =<br>1,135<sup>1</sup> |
| --- | --- |
| <sup>1</sup>n (%) |  |

### Decision-Making and Procedure Complications

Most notably, mothers (45%) and grandmothers (41%) largely decided to carry out uvulectomy. Moreover, most procedures were carried out by traditional healers (55%) as presented in Table 4. In addition, flat irons (25%) and hooked nails (19%) were the most used instruments for uvulectomy.

**Table 4:** Who decided and performed uvulectomy among the participants.

| Characteristic | N = 165<sup>1</sup> |
| --- | --- |
| **Who decided the procedure?** |  |
| - Mother | 74 (45%) |
| - Grandmother | 67 (41%) |
| - Husband | 58 (35%) |
| - Grandfather | 48 (29%) |
| - Husband's relatives | 20 (12%) |
| - Mother's relatives | 15<br>(9.1%) |
| - Friend | 15<br>(9.1%) |
| - Other | 5 (3.0%) |
| **Who performed the procedure?** |  |
| - Traditional healer (ie, Fakky or Baseer) | 90 (55%) |
| - Mother or relatives | 32 (19%) |
| - Older women | 29 (18%) |
| - Medical assistant | 10<br>(6.1%) |
| - Other | 9 (5.5%) |
| <sup>1</sup>n (%) |  |

**Table 5:** Methods, medications, and complications of the performed uvulectomy among the study participants.

| **Characteristic** | **N = 165<sup>1</sup>** |
| --- | --- |
| **Methods used** |  |
| - Flat iron | 41 (25%) |
| - Nail Hooked | 32 (19%) |
| - Bar Razor Blade | 25 (15%) |
| - Dental Instrument | 23 (14%) |
| - Don't know | 46 (28%) |
| - Other | 2 (1.2%) |
| **If another method, please specify** |  |
| String | 1 (50%) |
| Hand | 1 (50%) |
| **Medications used** |  |
| - Acacia (ie. Qarad) | 72 (44%) |
| - Rose geraniums (ie, a'atroon) | 34 (21%) |
| - Other | 21 (13%) |
| - None | 45 (27%) |
| **How soon after uvulectomy did you breastfeed/feed your child?** |  |
| Immediately | 78 (49%) |
| Later that day | 53 (33%) |
| The next day | 16 (10%) |
| The day after next | 4 (2.5%) |
| More than three days post-operation | 9 (5.6%) |
| **Complications reported** |  |
| - **Fever** | 58 (35%) |
| - **Difficulty in feeding** | 35 (21%) |
| - **Bleeding** | 22 (13%) |
| - **Secondary infection** | 8 (4.8%) |
| - **Tetanus** | 2 (1.2%) |
| - **Other** | 7 (4.2%) |
| <sup>1</sup>n (%) |  |

### Associations

Younger mothers (median age 22 vs. 28; p<0.001) and those with lower education (21% illiterate in the uvulectomy group vs. 7.4% in non-practicing; p<0.001) to carry out uvulectomy. Significantly, tribal tradition strongly influenced uvulectomy (92% in practicing groups compared to 25%; p<0.001), and cultural norms and tribal practice were strongly particular among young, poor, educated mothers (Table 6)

**Table 6:** The association between demographic characteristics and traditional uvulectomy practice.

| **Characteristic** | **No**<br>**N = 958<sup>1</sup>** | **Yes**<br>**N = 165<sup>1</sup>** | **p-**<br>**value<sup>2</sup>** |
| --- | --- | --- | --- |
| **Mother Age** | 28 (17, 51) | 22 (22, 33) | **<0.001** |
| **Mother education** |  |  | **<0.001** |
| Illiterate | 71 (7.4%) | 34 (21%) |  |
| Informal | 35 (3.7%) | 51 (31%) |  |
| Primary | 151 (16%) | 41 (25%) |  |
| Secondary | 206 (22%) | 25 (15%) |  |
| Graduate | 411 (43%) | 13 (7.9%) |  |
| Postgraduate | 84 (8.8%) | 1 (0.6%) |  |
| **Age of your oldest child** | 11 (6, 20) | 16 (10, 25) | **<0.001** |
| **How old were you when you give birth to your oldest child?** | 22.0 (19.0, 26.0) | 20.0 (17.0, 22.0) | **<0.001** |
| **How many children do you have?** | 3.00 (2.00, 5.00) | 4.00 (3.00, 6.00) | **<0.001** |
| **Age of your youngest child** |  |  | **0.048** |
| < 1 year | 40 (4.2%) | 10 (6.1%) |  |
| 1-3 years | 333 (35%) | 64 (39%) |  |
| 3-6 years | 234 (24%) | 41 (25%) |  |
| 6-12 years | 199 (21%) | 19 (12%) |  |
| 12-18 years | 152 (16%) | 31 (19%) |  |
| **Sex of the youngest child** |  |  | **<0.001** |
| Male | 482 (50%) | 111 (67%) |  |
| Female | 476 (50%) | 54 (33%) |  |
| **Monthly income of the family** |  |  | 0.4 |
| Low income | 298 (31%) | 60 (36%) |  |
| Moderate income | 585 (62%) | 95 (58%) |  |
| High income | 68 (7.2%) | 10 (6.1%) |  |
| **Mother work** | 271 (28%) | 68 (41%) | **<0.001** |
| **The monthly income of the mother** |  |  | >0.9 |
| Low income | 145 (39%) | 34 (38%) |  |
| Moderate income | 212 (57%) | 52 (58%) |  |
| High income | 14 (3.8%) | 4 (4.4%) |  |
| **Will you schedule your child for uvulectomy?** | 33 (3.5%) | 124 (84%) | **<0.001** |
| **Presence of uvulectomy practice in your tribe** | 213 (25%) | 151 (92%) | **<0.001** |

<sup>1</sup>Median (Q1, Q3); n (%)
<sup>2</sup>Wilcoxon rank sum test; Pearson's Chi-squared test; Fisher's exact test

## Discussion

Traditional uvulectomy remains a significant public health challenge in Sudan, with a 15% prevalence rate reported among children and adolescents in our study. This situates Sudan within the broader sub-Saharan African context, where the practice endures despite considerable regional variation. Our prevalence rate places Sudan between Tanzania’s low prevalence of 3.6% [25] and Khartoum’s 17.9% [17], as well as significantly higher rates in Southwest Ethiopia (61.9%) [7], Arbaminch, Ethiopia (36.6%) [26], Ekiti, Nigeria (26.9%) [27], and Debre Birhan, Ethiopia (23.7%) [28]. These regional differences likely arise from complex interactions between belief systems and cultural practices, differential access to modern health facilities, and disparities in educational and economic progress.

Beyond overall prevalence, our study identified a typical bimodal age distribution, with the highest prevalence among infants under six years (16%) and adolescents between 12 and 18 years (17%). This distribution reveals that the practice serves different sociocultural functions at various stages of development. The persistence of traditional uvulectomy has a strong association with cultural beliefs and perceived advantages of therapy. In our study, Mothers primarily offered breastfeeding difficulties (18%), cultural traditions (16%), and failure to thrive (13%) as justifications for this practice, which aligns with Ethiopia and Nigeria studies citing respiratory illness and breastfeeding difficulties [7,17,23,29]. Notably, 59.9% were influenced by traditional beliefs [29]. However, the knowledge gaps exist; while 30% of mothers believe the procedure is effective, 36% have no idea, and 24% fear harm if the uvula remains intact. These misconceptions are also reported in Tanzania, where 90% attributed sickness to uvula size [30], while 64.7% of Nigerians perceived the uvula as causing illness [28].

Despite these strongly held beliefs, Our findings revealed critical knowledge gaps regarding the risks and understanding of the procedure, 34% of mothers rejected traditional uvulectomy as ineffective, only, but 26% believed that uvula can lead to obstruction of the airway, and 37% were uncertain, Such uncertainty, together with fears about harm due to the uvula (24%), this lower than the finding of Linto et al whose the majority (68.3%) of the respondents believed TU is useful because it relieves vomiting and swallowing problem [31], point to the need for targeted health education. Family decision-making styles categorized mothers (45%) and grandmothers (41%) as principal initiators, with traditional healers performing 55% of procedures, that much lower than Nigeria’s study 76.5% [18], this reflects intergenerational transmission of beliefs, also seen in Eritrea and Tanzania, where family elders heavily influence healthcare choices [25,31]

The sociodemographic characteristics of these decision makers can help in explaining these patterns, with ages 18 to 52 years, and a median 33. Among the respondents, 35% held a university degree, 21% completed secondary education, 18% attained primary education, and 10% were illiterate. These findings align with a study in Gondar Town, Ethiopia, which reported an 11.6% paternal illiteracy rate, 38.7% completion of grade 12, and only 3.6% of mothers attend college education [7]. In contrast, a study in Sokoto State, Nigeria, noted that 43.8% of participants received Quranic education, while only 2.8% lacked formal education.[29]. These findings are similar to those of Linto et al, who reported that only a few of the respondents had never been to school and reported most were unemployed [31], 70% unemployed in our study.

Practice was strongly socio-demographically structured, being significantly associated with younger maternal age (median 22), lower education (21% illiterate), higher parity, and adherence to tribal tradition (92% agreement), all p<0.001. These patterns are consistent with wider patterns relating healthcare disparities to the use of traditional medicine [20,23,32]. These align with Isa et al.’s study, which linked higher maternal education to reduced attribution of diseases to the uvula.[20] However, this pattern didn’t hold in Eritrea, where education did not similarly influence TU practices. [31]

Traditional uvulectomies in these settings typically involve non-specialists, such as healers or lay practitioners, using unsterilized tools like flat irons (25%), nail hooks (18%), and razor blades (14%). Similar findings emerged from a study in Gondor, Ethiopia, and Uganda. [3, 26] underscores the widespread use of this practice across Africa. Socio-cultural beliefs and economic factors strongly influence its persistence in these communities [20], reflecting broader regional health challenges tied to accessibility and cultural norms.

A systematic review in Africa highlights that unsterilized tools primarily drive many complications [32].The use of unsafe techniques inevitably leads to severe complications, including life-threatening hemorrhages that lead to severe anemia, as observed in neonates [33]. Bleeding emerged as the most frequently reported complication across studies [16,25,34]. In our study, 12% of patients required urgent hospitalization for hemorrhage, significantly lower than rates reported in Nigeria (85.7% in Sokoto and 14.7% in Ekiti [29]. Fever occurred in 34% of cases, while 21% experienced feeding difficulties, closely aligning with Ekiti’s findings (28%) [29]. Both studies reported identical infection rates (2.9%), highlighting the serious risks associated with TU. Although septicemia poses a life-threatening risk following traditional uvulectomy, it occurs infrequently [23]. Practitioners commonly apply traditional remedies such as acacia and rose geranium during uvulectomy. In contrast, a study at Danfodiyo University Teaching Hospital in Sokoto, Nigeria, found that traditional healers administered herbal medications to 33.3% of patients to manage post-procedural bleeding [29]. Alarmingly, 57% of mothers are unaware of these complications, and 49% of mothers persisted with breastfeeding immediately, an indicator of normalization of these risks.

Contrary to studies in Tanzania where uvulectomy decreased with access to healthcare, our urban sample (22% from Khartoum) still reported high practice, reflecting persistent traditional beliefs despite medical access [30]. This is comparable to findings in Dar es Salaam, where caregivers preferred uvulectomy after failed clinical interventions [36], suggesting deep cultural entrenchment. Given that traditional uvulectomy intersects with critical human rights violations and public health risks, eradicating these harmful practices requires coordinated, multisectoral interventions.

## Conclusion

Our cross-sectional community-based study revealed that traditional uvulectomy continues to be a culturally embedded practice among Sudanese children and adolescents, facilitated by intergenerational power, maternal beliefs of the therapeutic need, and tribal traditions. The practice is strongly associated with younger maternal age, lower education levels, and tribal affiliations, and is dominated by decision-making by mothers and grandmothers. Procedural complications are compounded by the use of non-sterilized tools and reliance on traditional practitioners, leading to reported complications such as fever, feeding difficulties, and bleeding. Despite variations in maternal education, biomedical knowledge is frequently supplanted by cultural compliance, underscoring a severe disconnect between health knowledge and traditional health-seeking behavior.

To reduce unsafe traditional uvulectomy practices in Sudan, public health efforts should prioritize culturally sensitive interventions involving tribal leaders, grandmothers, and traditional healers as key partners. Public education must refute myths regarding the role of the uvula in child health. Concurrently, improve healthcare access and educate providers to deliver safer alternatives. Policymakers need to enact joined-up legislation lowering risks without isolating communities and informed by research on sociocultural drivers of the practice. By bringing together respect for tradition and evidence-informed child safety, Sudan can establish balanced healthcare that balances tradition and security.

### Study strengths and limitations

To the best of our knowledge, this is the first study that assess the Prevalence and practice of Traditional Uvulectomy Among Sudanese Children and Adolescents, Along with Maternal Perception included most the Sudanese states and This is one of the most comprehensive to assess the traditional uvulectomy in Sudan (N=1,135), that enhances the generalizability constituting strong evidence for practice and policy. Despite this, we acknowledge some limitations threat of recall bias in self-reports, Geographic heterogeneities that hinder generalizability at a national level, and the cross-sectional design limits causal conclusions

### Future directions

Further implementation science must compare the efficacy of culturally-adapted interventions to these methodological constraints. Qualitative research on the socio-cultural determinants of uvulectomy persistence can provide further evidence-based findings for intervention development.

## Data Availability

All data produced in the present study are available upon reasonable request to the authors

## Author Contributions

### Conceptualization

Jaber Hamad Jaber Amin

### Data curation

Jaber Hamad Jaber Amin, Ahmed Alshafei Elmahi Ahmed, Alshafee Mohammed Jebreel Sulyman,

### Formal analysis

Mohamedelmustafa Yahya Mohamed Eldouma

### Investigation

Amna A. Eltayeb, Asjad Zainalabdeen Ahmed, Huyam Elsir Fadlelseed Babiker

### Methodology

Tanzeel Mohamedain Abuelgasim Abdalla, Tanzeel Mohamedain Abuelgasim Abdalla, Dania A. Elsiddig.

### Resources

Majdy Jailany Alamin Abdelgadir, Mohammed Hammad Jaber Amin

### Software

Nooralsham Abdalla Adam Yousif, Majdy Jailany Alamin Abdelgadir, Israa Osman Ali Alhassan, Zainab Salih Ahmed Adam.

### Validation

Jaber Hamad Jaber Amin, Mohamedelmustafa Yahya Mohamed Eldouma, Ahmed Alshafei Elmahi Ahmed, Mohammed Hammad Jaber Amin

### Visualization

Alshafee Mohammed Jebreel Sulyman, Amna A. Eltayeb, Asjad Zainalabdeen Ahmed.

### Writing – original draft

Ahmed Alshafei Elmahi Ahmed, Mohamedelmustafa Yahya Mohamed Eldouma, Alshafee Mohammed Jebreel Sulyman, Amna A. Eltayeb, Asjad Zainalabdeen Ahmed, Huyam Elsir Fadlelseed Babiker, Tanzeel Mohamedain Abuelgasim Abdalla, Israa Osman Ali Alhassan, Dania A. Elsiddig, Nooralsham Abdalla Adam Yousif, Majdy Jailany Alamin Abdelgadir, Zainab Salih Ahmed Adam.

### Writing – review & editing

Jaber Hamad Jaber Amin, Ahmed Alshafei Elmahi Ahmed, Mohamedelmustafa Yahya Mohamed Eldouma, Mohammed Hammad Jaber Amin

